# Prognostic value of the Hippo signaling pathway in oral squamous cell carcinoma: a systematic review and meta-analysis

**DOI:** 10.64898/2026.10.09.26365171

**Authors:** Igor Felipe Pereira Lima, Leonardo da Silva Bittencourt, Petra Gonçalves Reinehr, Adam Jeffrey Engler, Marcelo Lazzaron Lamers

**Affiliations:** Department of Oral Pathology, School of Dentistry, Federal University of Rio Grande do Sul, Porto Alegre, Rio Grande do Sul, Brazil; State School of Professional Health Education Hospital de Clínicas de Porto Alegre, Porto Alegre, Rio Grande do Sul, Brazil; Postgraduate Program in Physiology, Institute of Basic Health Sciences, Federal University of Rio Grande do Sul, Porto Alegre, Rio Grande do Sul, Brazil; Chien-Lay Department of Bioengineering, UC San Diego, San Diego, California, USA; Department of Morphological Sciences, Institute of Basic Health Sciences, Federal University of Rio Grande do Sul, Porto Alegre, Rio Grande do Sul, Brazil

**Keywords:** Oral squamous cell carcinoma, Hippo signaling pathway, YAP1 protein, WWTR1 protein, Prognosis

## Abstract

**Background:** This systematic review and meta-analysis evaluated the prognostic value of Hippo signaling pathway biomarkers in oral squamous cell carcinoma.

**Methods:** Following the Preferred Reporting Items for Systematic Reviews and Meta-Analyses guidelines, electronic database searches identified 17 eligible studies involving 1,194 patients. Associations between the immunohistochemical expression of Hippo pathway effectors, primarily YAP and TAZ, and clinicopathological characteristics were assessed. Meta-analyses using random-effects models estimated pooled hazard ratios for overall survival.

**Results:** Predominantly nuclear expression of YAP and TAZ was consistently associated with adverse clinicopathological features, including advanced clinical stage, lymph node metastasis, and poorer histological differentiation. Meta-analysis demonstrated that YAP overexpression was associated with worse overall survival (hazard ratio, 1.918; 95% confidence interval, 1.036 to 3.549; p = 0.038). TAZ overexpression was similarly associated with poorer overall survival (hazard ratio, 2.845; 95% confidence interval, 1.682 to 4.811; p < 0.001). Although substantial between-study heterogeneity was observed, the direction of the prognostic association remained consistent across studies.

**Conclusions:** Increased nuclear expression of YAP and TAZ is consistently associated with more aggressive disease and poorer overall survival in oral squamous cell carcinoma. These findings support the prognostic value of Hippo pathway biomarkers, although methodological heterogeneity warrants cautious interpretation.

Graphical abstract
Nuclear overexpression of the Hippo pathway effectors YAP and TAZ is associated with adverse clinicopathological features and poorer overall survival in oral squamous cell carcinoma: a systematic review and meta-analysis of 17 studies comprising 1,194 patients.

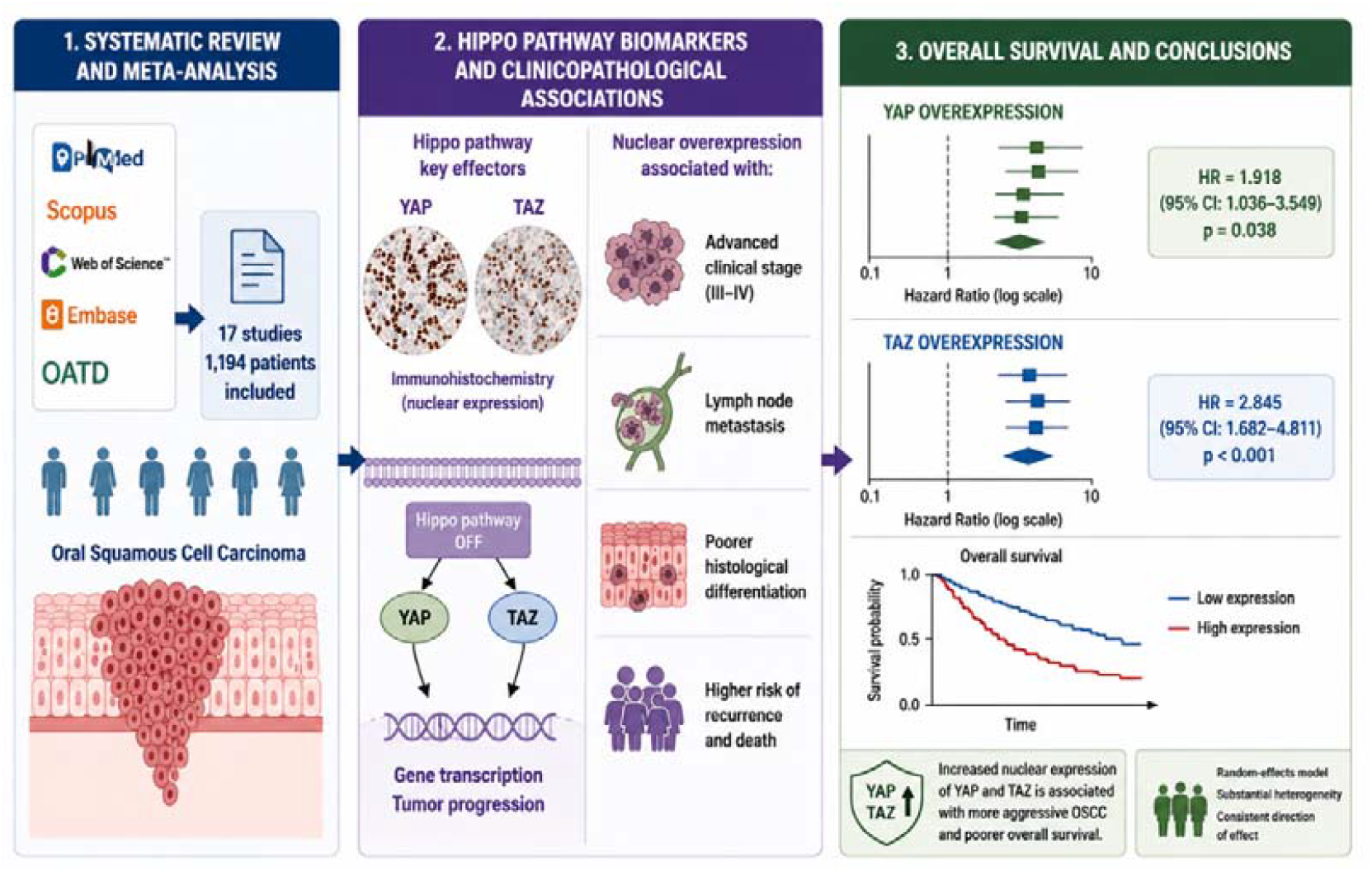

## BACKGROUND

With origin from the stratified squamous epithelium that lines the oral mucosa, Oral Squamous Cell Carcinoma (OSCC) is the most prevalent malignant neoplasm of the oral cavity [1]. OSCC is characterized by aggressive local invasion and frequent regional lymph node metastasis [2,3]. Despite advances in therapeutic modalities, including surgery, radiotherapy, and chemotherapy, OSCC remains associated with an unfavorable prognosis, with a five-year OS rate below 50% in recent decades [4,5]. High treatment-related morbidity and frequent locoregional recurrence further emphasize the need to identify molecular determinants of tumor progression and prognosis [6,7].

OSCC represents a global public health challenge, ranking among the ten most common malignant neoplasms today, with more than 377,000 new cases and approximately 177,000 deaths recorded every year [8,9]. The incidence shows significant geographic variation, with particularly high rates in South and Southeast Asia, where it accounts for up to 40% of all malignant neoplasms, in contrast to rates below 4% in developed regions such as North America and Western Europe [10,11]. The main challenges include late diagnosis, observed in approximately 60% of cases that present at advanced stages (III–IV), therapeutic resistance, the absence of reliable predictive biomarkers for risk stratification, and the limited effectiveness of available targeted therapies [12–14].

Recent studies have shown that tumor progression is influenced by several factors, including mechanical signals originating from the tumor microenvironment [15,16]. Neoplastic cells reside in a dynamic microenvironment characterized by altered stiffness of the extracellular matrix (ECM), compressive forces, and ECM remodeling, factors that influence cell behavior through specific signaling pathways [17,18]. Mechanotransduction has been implicated in the activation of pro-oncogenic transcriptional programs, epithelial–mesenchymal transition (EMT), and maintenance of cancer stem cells, processes that are fundamental to tumor progression and metastasis [19–21]. Among mechanotransduction pathways, Hippo signaling has emerged as a central regulator of these processes [22,23]. The nuclear effectors of this pathway, the proteins YAP and TAZ (Transcriptional coactivator with PDZ-binding motif), function as transcriptional coactivators that, when dysregulated, promote aberrant expression of genes involved in cell proliferation, survival, and migration [24,25].

Although experimental evidence supports the involvement of Hippo signaling in oral carcinogenesis, its prognostic significance in oral squamous cell carcinoma remains uncertain [26,27]. Therefore, this systematic review evaluated the association between the immunohistochemical expression of Hippo pathway biomarkers (YAP, TAZ, MST1/2, LATS1/2, and SAV1) and clinicopathological and prognostic outcomes in oral squamous cell carcinoma. The null hypothesis was that Hippo pathway biomarker expression is not associated with prognosis in oral squamous cell carcinoma.

## METHODS

### Protocol and registration

This systematic review was conducted following the Preferred Reporting Items for Systematic Reviews and Meta-Analyses (PRISMA) [28] statement and Cochrane guidelines. The review protocol was registered in the PROSPERO database CRD420251231277.

### Study design

This is a systematic review designed to answer the following guiding PECO question: “In patients with oral squamous cell carcinoma (Population), is high expression of Hippo pathway biomarkers (Exposure), compared with low expression (Comparison), associated with worse tumor progression or clinical prognosis (Outcome)?”

### Eligibility criteria

Studies that evaluated the expression of Hippo pathway biomarkers (YAP, TAZ, MST1/2, LATS1/2, SAV1, and related molecules) in OSCC and assessed tumor progression or clinical outcomes, with no restrictions on year, language, or publication status (including those in press), were included.

The following studies were excluded: studies focused on other cancer types without specific data for OSCC; review, editorials, letters to the editor, or conference abstracts without complete data; studies evaluating only molecular aspects of the Hippo pathway without analysis of expression in tumor tissue or without correlation with prognosis; studies in which OSCC data could not be dissociated from other head and neck cancers; in vitro studies; and animal studies.

### Sources of information and search

Two reviewers (IFPL and PGR) independently searched electronic databases. Descriptors were identified using DeCS (Health Sciences Descriptors), MeSH (Medical Subject Headings), and Emtree (Embase Subject Headings), and the Boolean operators “AND” and “OR” were used to optimize the search strategy (Table 1). The results were exported to EndNote Basic/Online (Thomson Reuters, Toronto, Canada), and duplicates were removed.

**Table 1.** Search strategies in the databases.

| **Databases** | **Search strategies (November, 2025)** | **Results** |
| --- | --- | --- |
| **PubMed**<br>http://www.ncbi.nlm.nih.gov/pubmed | ("Hippo" OR "YAP" OR "TAZ" OR "mechanotransduction" OR "mechanosignaling") AND ("Mouth Neoplasms" OR "head and neck cancer" OR "Oral cancer" OR "Oral squamous cell carcinoma" OR "Oral cavity neoplasms" OR "Mouth neoplasms") AND ("Prognosis" OR "Survival" OR "Survival rate" OR "Recurrence" OR "Disease progression") | 99 |
| **Scopus**<br>https://www.scopus.com/ | ("Hippo" OR "YAP" OR "TAZ" OR "mechanotransduction" OR "mechanosignaling") AND ("Mouth Neoplasms" OR "head and neck cancer" OR "Oral cancer" OR "Oral squamous cell carcinoma" OR "Oral cavity neoplasms" OR "Mouth neoplasms") AND ("Prognosis" OR "Survival" OR "Survival rate" OR "Recurrence" OR "Disease progression") | 110 |
| **Embase**<br>https://www.embase.com/ | ('hippo' OR 'yap' OR 'taz' OR 'mechanotransduction'/exp OR 'mechanotransduction' OR 'mechanosignaling'/exp OR 'mechanosignaling') AND ('head and neck cancer'/exp OR 'head and neck cancer' OR 'oral cancer'/exp OR 'oral cancer' OR 'oral squamous cell carcinoma'/exp OR 'oral squamous cell carcinoma' OR 'oral cavity neoplasms' OR 'mouth neoplasms'/exp OR 'mouth neoplasms') AND ('prognosis'/exp OR 'prognosis' OR 'survival'/exp OR 'survival' OR 'survival rate'/exp OR 'survival rate' OR 'recurrence'/exp OR 'recurrence' OR 'disease progression'/exp OR 'disease progression') | 474 |
| **Web Of Science**<br>http://apps.webofknowledge.com/ | ("Hippo" OR "YAP" OR "TAZ" OR "mechanotransduction" OR "mechanosignaling") AND ("Mouth Neoplasms" OR "head and neck cancer" OR "Oral cancer" OR "Oral squamous cell carcinoma" OR "Oral cavity neoplasms" OR "Mouth neoplasms") AND ("Prognosis" OR "Survival" OR "Survival rate" OR "Recurrence" OR "Disease progression") | 93 |
| **OATD**<br>https://oatd.org/ | ("Hippo" OR "YAP" OR "TAZ" OR "mechanotransduction" OR "mechanosignaling") AND ("Mouth Neoplasms" OR "head and neck cancer" OR "Oral cancer" OR "Oral squamous cell carcinoma" OR "Oral cavity neoplasms" OR "Mouth neoplasms") AND ("Prognosis" OR "Survival" OR "Survival rate" OR "Recurrence" OR "Disease progression") | 4 |
| **TOTAL** |  | 780 |

### Study selection

The selection of studies was conducted in three distinct stages. In the first stage, two reviewers (IFPL and PGR) systematically screened the titles of all retrieved records. Articles with titles relevant to the objective of this systematic review were selected for the second stage. In the second stage, the same reviewers screened the abstracts and applied the inclusion and exclusion criteria. Studies without available abstracts were evaluated in the third stage. In this final stage, the full texts of the articles were assessed. When disagreements arose regarding a study’s eligibility, a third reviewer (MLL) was consulted to make a final decision. Studies that did not meet the criteria were recorded separately, with the reasons for exclusion clearly documented.

### Data collection and extraction

Two authors (IFPL and PGR) performed data extraction using specific spreadsheets designed for this purpose. The following information was extracted from the eligible studies: author and year, sample size (n), sex, age (mean), tumor location, type of tumor, biomarker, analysis method, prognosis and survival analysis, expression of biomarker, subcellular location, clinicopathological correlation, survival analysis and prognosis analysis.

As a calibration exercise, the reviewers (IFPL and PGR) discussed the eligibility criteria and applied them to a random sample of 20% of the retrieved studies to assess inter-rater agreement. After reaching an adequate level of agreement (κ = 0.81–0.84), the reviewers independently extracted data from all studies. Any discrepancies were resolved by consensus and supervision of a third reviewer (MLL). Authors of studies with insufficient data or missing information necessary for summarization and comparison were contacted via email to request additional details.

### Risk of bias of individual studies

The risk of bias for individual studies was independently assessed by two authors (IFPL and PGR) using the “*Joanna Briggs Institute Critical Appraisal Checklist for Analytical Cross Sectional Studies*” and “*The Joanna Briggs Institute Critical Appraisal Checklist for Cohort Studies*”. The risk of bias was classified as **“High”** when the study achieved up to 49% of “yes” responses, **“*Moderate”*** when it ranged from 50% to 69%, and **“*Low”*** when it exceeded 70% (Table 3).

### Statistical analysis

Data from eleven included studies were integrated and analyzed using Comprehensive Meta-Analysis (CMA) software, Version 4 (Biostat, Inc., Englewood, NJ, USA; available at meta-analysis.com).

The prognostic value of YAP and TAZ expression was evaluated by pooling HR and their 95% Confidence Intervals (CI). HRs were extracted directly from the articles or estimated from Kaplan-Meier survival curves using the software Webplotdigitizer Version 5 (Rohatgi, A. (2025). WebPlotDigitizer: Version 5.2. https://automeris.io), when necessary.

Heterogeneity among studies was assessed using the Chi-square test and quantified by the I^2^ statistic. An I^2^ value greater than 50% was considered indicative of substantial heterogeneity. Consequently, a random-effects model (DerSimonian and Laird method) was employed to calculate the pooled HR to account for the variation in study populations and methodologies (specifically IHC protocols). Publication bias was evaluated through visual inspection of funnel plots for asymmetry. A p-value of <0.05 was considered statistically significant.

## RESULTS

### Selection of studies

The search of the PubMed, Web of Science, Embase, Scopus, and OATD databases yielded 780 articles. After removal of duplicates, 523 records remained for title screening, of which 108 were considered potentially eligible for abstract review. Following abstract screening, 19 articles were selected for full-text assessment. In addition, all references cited in the selected manuscripts (a total of 822 references) were carefully examined to identify potential studies not captured by the primary search strategy. After the exclusion of 2 articles, one for not presenting OSCC data separately for analysis [30] and another due to incomplete data for YAP/TAZ analysis [31], 17 studies were included in this systematic review (Figure 1).

**Fig. 1.**
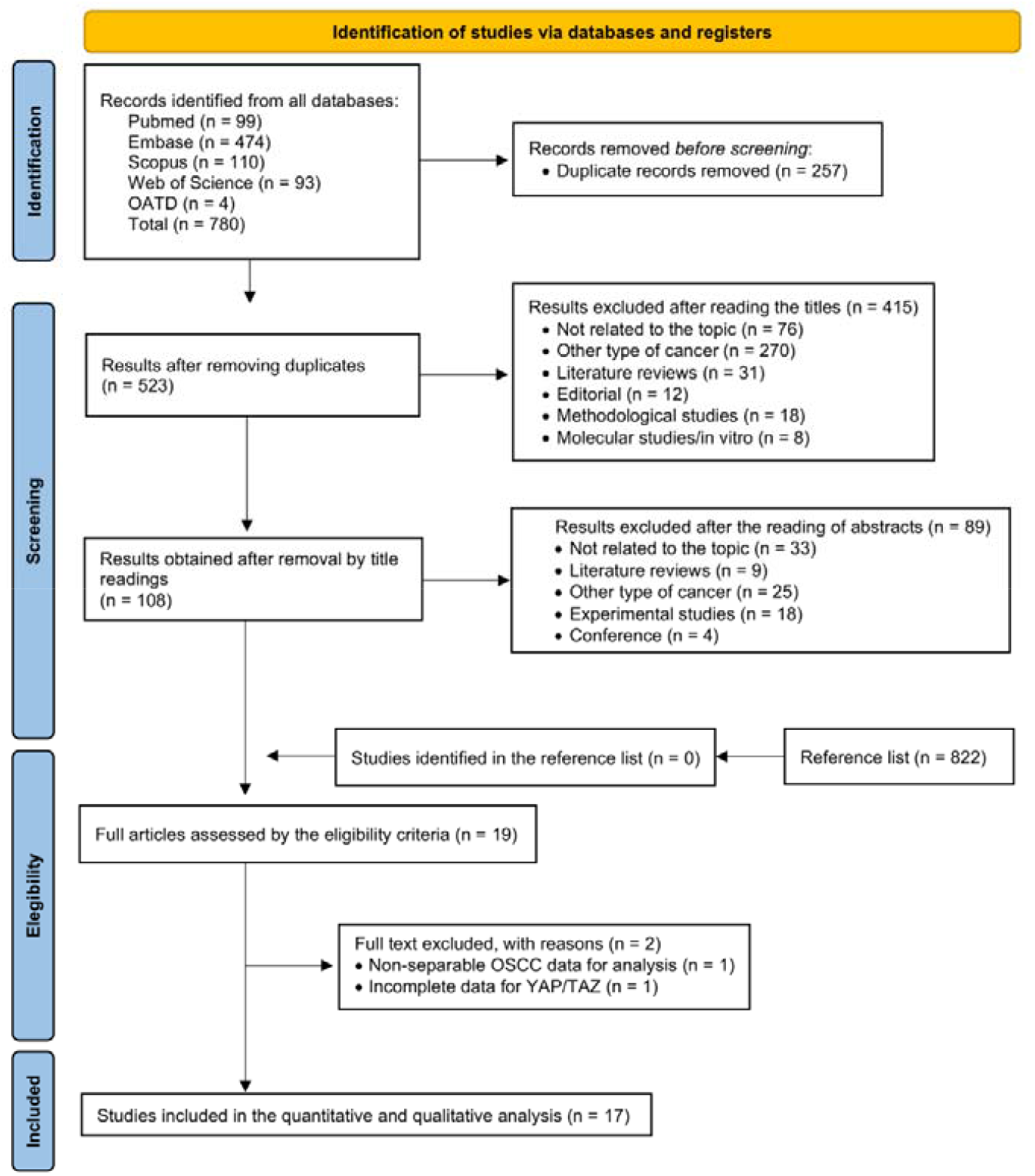
PRISMA Flow Diagram for Systematic Review and Meta-Analysis of the prognostic significance of Hippo pathway biomarkers in OSCC.

### Study characteristics

The 17 included articles were published between 2013 and 2026, and all studies were conducted in patients with OSCC [32–48]. The total sample comprised 1,194 patients, with individual sample sizes ranging from 9 to 130 participants. Where reported, there was a predominance of males in most studies, whereas only one study [46] described a slight predominance of females. The mean age of participants ranged approximately from 50.76 to 68.21 years, while other studies reported age only in categorical form, such as <50 vs. ≥50 years [37] or ≤60 vs. >60 years [40].

With regard to the anatomical location of the tumors, the tongue was the most frequently investigated site, being the exclusive focus in four studies [32,37,39,44] and included in others within samples comprising multiple intraoral sites [32–41,43–48]. Other sites evaluated included the oral mucosa, floor of the mouth, gingiva, retromolar region, maxillary and mandibular alveolar ridge, hard palate, lip, retrocommissural region, and the oral cavity more broadly. In all studies [32–48], the lesions evaluated were malignant and classified as OSCC, with no benign lesions or other histological tumor types being included.

Most studies focused on Hippo pathway core effectors, e.g., YAP/YAP1 [33,35–41,43–48] and TAZ [32–35,40,43–45], and they assessed either individually or in combination. Some studies additionally evaluated markers that are upstream, downstream, or functionally related to the Hippo pathway, such as p-YAP1 [40], MST1 [46], MST2 [46], LATS1 [46], LATS2 [46], PIEZO1 [43], GPR39 [41], SOX2 [42] and Kank1 [37], thereby exploring broader molecular panels. In all studies [32–48], biomarker expression was analyzed by immunohistochemistry on tumor samples, allowing quantification of expression in neoplastic tissue and subsequent stratification of patients into high-versus low-expression groups for prognostic purposes. Beyond marker expression, 13 [32,34–41,44–47] of the 17 studies constructed Kaplan–Meier survival curves to compare outcomes between patients with high and low expression of Hippo pathway biomarkers, and six of these [32,34,35,44,46,47] further complemented this approach with Cox proportional hazards regression models to estimate the independent prognostic impact after adjustment for clinicopathological variables. In the remaining studies [33,42,43,48], prognosis was inferred indirectly through correlations between biomarker expression and adverse clinicopathological features (such as more advanced tumor stage or higher histological grade) or evidence of lesion progression, without the formal presentation of survival curves (Table 2).

**Table 2.** Summary of the main characteristics of the eligible studies.

| Author and year | Sample (n) | Sex | Age (mean) | Tumor location | Type of tumor | Biomarker | Analysis method | Prognosis and survival analysis |
| --- | --- | --- | --- | --- | --- | --- | --- | --- |
| Wei et al., 2013[32] | 52 | 28 (male)<br>24 (female) | 53.5 | Tongue | OSCC | TAZ | IHC | Kaplan-Meier and Cox regression model of proportional hazards |
| Hiemer et al., 2015[33] | 10 | -- | -- | Tongue and floor of the mouth | OSCC | YAP (YAP1) and TAZ (WWTR1) | IHC | The prognosis was inferred by correlating it with advanced tumor grade and stage |
| Li et al., 2015 [34] | 111 | -- | -- | Tongue, oral mucosa, floor of the mouth, and gingiva | OSCC | TAZ | IHC | Kaplan-Meier and Cox regression model of proportional hazards |
| Szelachowska et al., 2019 [35] | 127 | 102 (male)<br>25 (female) | -- | Tongue and floor of the mouth | OSCC | YAP (YAP1) and TAZ (WWTR1) | IHC | Kaplan-Meier and Cox regression model of proportional hazards |
| Ono et al., 2019 [36] | 80 | -- | -- | Oral cavity | OSCC | YAP | IHC | Kaplan-Meier |
| Fan et al., 2019 [37] | 32 | -- | <50 (n=15)<br>>50 (n=17) | Tongue | OSCC | YAP and Kank1 | IHC | Kaplan-Meier |
| Qi et al., 2019 [38] | 68 | 45 (male)<br>23 (female) | 55.5 | Tongue, oral mucosa, retromolar region, gingiva and floor of the mouth | OSCC | YAP | IHC | Kaplan-Meier |
| Omori et al., 2020 [39] | 86 | -- | -- | Tongue | OSCC | YAP (YAP1) | IHC | Kaplan-Meier |
| Peng et al., 2020 [40] | 56 | 34 (male)<br>22 (female) | ≤60 (n=20)<br>>60 (n=36) | Tongue and oral cavity | OSCC | YAP1, p-YAP1, TAZ and p-TAZ | IHC | Kaplan-Meier |
| Jiang et al., 2020 [41] | 122 | -- | -- | Alveolar crest, tongue, oral mucosa, floor of the mouth, hard palate, lip, oral cavity | OSCC | GPR39 and YAP | IHC | Kaplan-Meier |
| Stasikowska-Kanicka et al., 2021 [42] | 86 | 53 (male)<br>33 (female) | 63.77 (M+)<br>68.21 (M-) | Floor of the mouth | OSCC | TAZ, SOX2 | IHC | Clinical-pathological correlation of M+ and M- |
| Hasegawa et al., 2021 [43] | 45 | -- | -- | Oral cavity | OSCC | YAP, TAZ and PIEZO1 | IHC | Clinical-pathological correlation |
| Yang et al., 2023 [44] | 129 | 69 (male)<br>60 (female) | -- | Tongue | OSCC | YAP and TAZ | IHC | Kaplan-Meier and Cox regression model of proportional hazards |
| Tsai et al., 2024 [45] | 21 | -- | -- | Oral cavity | OSCC | YAP and TAZ | IHC | Kaplan-Meier |
| Amano et al., 2024 [46] | 130 | 60 (male)<br>70 (female) | 63.9 | Tongue, gingiva, floor of the mouth and oral mucosa | OSCC | MST1, MST2, LATS1, LATS2 and YAP1 | IHC | Kaplan-Meier and Cox regression model of proportional hazards |
| Gupta et al., 2025 [47] | 30 | 25 (male)<br>5 (female) | 50.76 | Oral mucosa, lip, floor of the mouth, tongue, alveolus maxillary, alveolus mandibular and retrocommissural region | OSCC | YAP | IHC | Kaplan-Meier and Cox regression model of proportional hazards |
| Nandhakumar et al., 2026 [48] | 9 | 7 (male)<br>2 (female) | 57.34 | Tongue, oral mucosa and gingiva | OSCC | YAP1 | IHC | Correlation between lesion expression and progression |

-- Not informed by the author; OSCC: Oral Squamous Cell Carcinoma; YAP: Yes-Associated Protein; TAZ: Transcriptional coactivator with PDZ-binding motif; IHC: Immunohistochemistry

### Risk of Bias of studies

Using the Joanna Briggs Institute Critical Appraisal Tools for cohort and analytical cross-sectional studies [29], 16 of the 17 articles were judged to have a low risk of bias, while one cohort study [45] was classified as having a moderate risk of bias (63.63% “yes” responses). No study was rated as high risk of bias (Table 3). All three analytical cross-sectional studies achieved 75% “yes” ratings but did not identify confounding factors or describe strategies to address them (Q5 and Q6). In the cohort studies, most domains related to exposure and outcome measurement and statistical analysis were adequately fulfilled; however, confounding factors (Q5) and issues concerning completeness of follow-up and management of losses (Q9 and Q10) were frequently rated as unclear, which may limit the internal validity of some estimates.

**Table 3.**
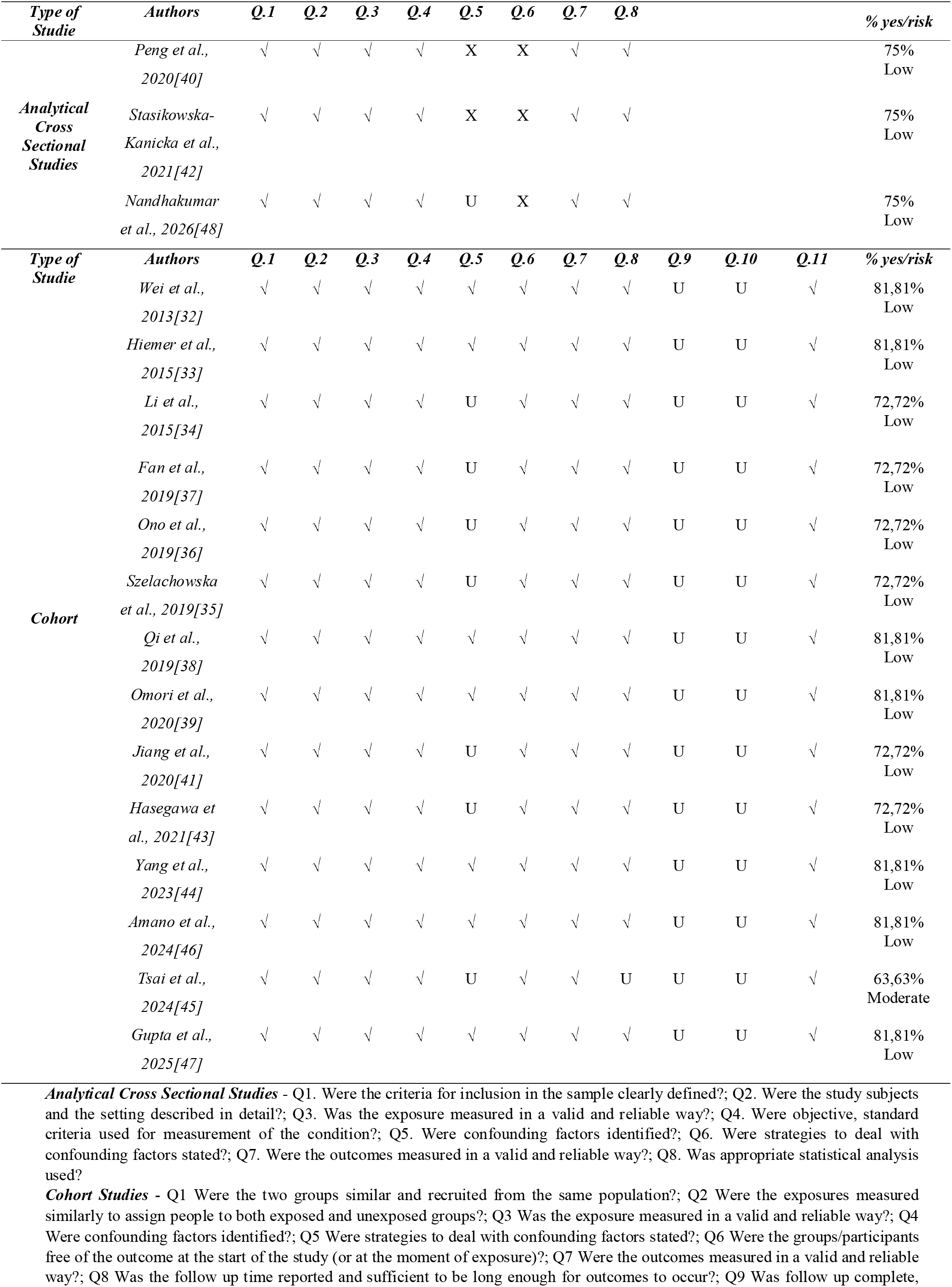
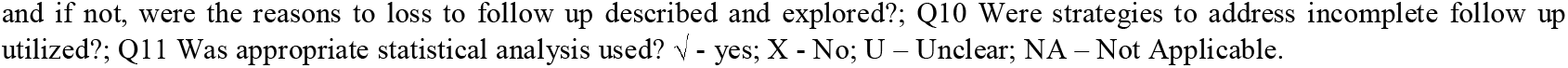
Risk of bias assessed by the Joanna Briggs Institute Critical Appraisal Tools for use in JBI Systematic Reviews for Cohort studies and Analytical Cross Sectional Studies.

| Type of Studie | Authors | Q.1 | Q.2 | Q.3 | Q.4 | Q.5 | Q.6 | Q.7 | Q.8 | % yes/risk |  |  |  |
| --- | --- | --- | --- | --- | --- | --- | --- | --- | --- | --- | --- | --- | --- |
| Analytical Cross Sectional Studies | Peng et al., 2020[40] | √ | √ | √ | √ | X | X | √ | √ | 75% Low |  |  |  |
|  | Stasikowska-Kanicka et al., 2021[42] | √ | √ | √ | √ | X | X | √ | √ | 75% Low |  |  |  |
|  | Nandhakumar et al., 2026[48] | √ | √ | √ | √ | U | X | √ | √ | 75% Low |  |  |  |
| Type of Studie | Authors | Q.1 | Q.2 | Q.3 | Q.4 | Q.5 | Q.6 | Q.7 | Q.8 | Q.9 | Q.10 | Q.11 | % yes/risk |
| Cohort | Wei et al., 2013[32] | √ | √ | √ | √ | √ | √ | √ | √ | U | U | √ | 81,81% Low |
|  | Hiemer et al., 2015[33] | √ | √ | √ | √ | √ | √ | √ | √ | U | U | √ | 81,81% Low |
|  | Li et al., 2015[34] | √ | √ | √ | √ | U | √ | √ | √ | U | U | √ | 72,72% Low |
|  | Fan et al., 2019[37] | √ | √ | √ | √ | U | √ | √ | √ | U | U | √ | 72,72% Low |
|  | Ono et al., 2019[36] | √ | √ | √ | √ | U | √ | √ | √ | U | U | √ | 72,72% Low |
|  | Szelachowska et al., 2019[35] | √ | √ | √ | √ | U | √ | √ | √ | U | U | √ | 72,72% Low |
|  | Qi et al., 2019[38] | √ | √ | √ | √ | √ | √ | √ | √ | U | U | √ | 81,81% Low |
|  | Omori et al., 2020[39] | √ | √ | √ | √ | √ | √ | √ | √ | U | U | √ | 81,81% Low |
|  | Jiang et al., 2020[41] | √ | √ | √ | √ | U | √ | √ | √ | U | U | √ | 72,72% Low |
|  | Hasegawa et al., 2021[43] | √ | √ | √ | √ | U | √ | √ | √ | U | U | √ | 72,72% Low |
|  | Yang et al., 2023[44] | √ | √ | √ | √ | √ | √ | √ | √ | U | U | √ | 81,81% Low |
|  | Amano et al., 2024[46] | √ | √ | √ | √ | √ | √ | √ | √ | U | U | √ | 81,81% Low |
|  | Tsai et al., 2024[45] | √ | √ | √ | √ | U | √ | √ | U | U | U | √ | 63,63% Moderate |
|  | Gupta et al., 2025[47] | √ | √ | √ | √ | √ | √ | √ | √ | U | U | √ | 81,81% Low |

**Analytical Cross Sectional Studies** - Q1. Were the criteria for inclusion in the sample clearly defined?; Q2. Were the study subjects and the setting described in detail?; Q3. Was the exposure measured in a valid and reliable way?; Q4. Were objective, standard criteria used for measurement of the condition?; Q5. Were confounding factors identified?; Q6. Were strategies to deal with confounding factors stated?; Q7. Were the outcomes measured in a valid and reliable way?; Q8. Was appropriate statistical analysis used?
**Cohort Studies** - Q1 Were the two groups similar and recruited from the same population?; Q2 Were the exposures measured similarly to assign people to both exposed and unexposed groups?; Q3 Was the exposure measured in a valid and reliable way?; Q4 Were confounding factors identified?; Q5 Were strategies to deal with confounding factors stated?; Q6 Were the groups/participants free of the outcome at the start of the study (or at the moment of exposure)?; Q7 Were the outcomes measured in a valid and reliable way?; Q8 Was the follow up time reported and sufficient to be long enough for outcomes to occur?; Q9 Was follow up complete,
and if not, were the reasons to loss to follow up described and explored?; Q10 Were strategies to address incomplete follow up utilized?; Q11 Was appropriate statistical analysis used? √ - yes; X - No; U – Unclear; NA – Not Applicable.

### Individual results of the studies

All 17 included studies analyzed the expression of Hippo pathway biomarkers, with most reporting high expression of YAP/YAP1 [32–33,35–39,41,43–48] and/or TAZ [32,34,35,42] in OSCC cases, particularly when compared with normal oral mucosa or potentially malignant lesions. Three studies [33,44,45] did not report TAZ expression data. One study [46] described reduced expression or loss of inhibitory components of the pathway (such as MST1/2 and LATS1/2), as well as the co-expression of functionally related markers, including PIEZO1 [43], GPR39 [41], SOX2 [42] and Kank1 [37]. Regarding subcellular localization, YAP/YAP1 and TAZ expression was predominantly nuclear or nucleo-cytoplasmic, and this was reported in 13 studies [32–36,38,39,41–43,46–48]. Only four studies [37,40,44,45] did not specify the subcellular localization of the staining.

With regard to clinicopathological correlations, 15 [32–33,36–43,44–48] of the 17 studies provided at least one association between Hippo pathway biomarker expression and features of tumor aggressiveness, whereas one study [44] did not report clinicopathological data and another focused solely on the correlation between YAP and TAZ expression, without clinical outcomes [35]. Overall, high expression of YAP/YAP1 and/or TAZ, particularly with nuclear localization, was associated with larger tumor size [34,41,47], more advanced clinical and/or pathological stage [32,33,37,41,45,47], higher histological grade [32,33,36,47,48] (with greater expression in moderately or poorly differentiated tumors), and, most notably, with lymph node involvement [34,38–43,45] and metastasis [39–43,45,46]. In addition, one study [41] related Hippo pathway biomarker expression to tumor recurrence, while another described an association with EMT features [46]. Hippo pathway–related markers, such as GPR39, SOX2, and TAZ/SOX2 coexpression, were also associated, in two studies [41,42], with metastasis or other adverse prognostic factors.

With respect to survival analysis, 12 of the 17 studies [32,34–36,38–41,44–47] performed some type of time-to-event analysis, generally using Kaplan–Meier curves to compare OSCC patients with high versus low expression of Hippo pathway biomarkers, whereas 5 studies [33,37,42,43,48] did not report or did not conduct survival analysis. Among the studies [32,34–36,38–41,44–47] that evaluated these outcomes, most showed that high expression of YAP/YAP1 and/or TAZ, particularly when nuclear, was associated with poorer OS [32,34,38–41,44–47] and/or shorter disease-free or recurrence-free survival [32,34–36,39,46], frequently with statistically significant differences between groups. Conversely, 4 studies [35,38,46,47] did not find a significant association between certain expression patterns and survival, especially when cytoplasmic fractions of YAP, the isolated expression of some upstream components, or TAZ expression without further stratification were analyzed.

Finally, in multivariable prognostic analysis, six studies [32,34,35,44,46,47] applied Cox regression models with hazard ratio estimates to test whether Hippo pathway biomarkers constituted independent risk factors after adjustment for classical clinicopathological variables (such as age, sex, tumor stage, histological grade, and nodal status). In three of these studies [32,34,35], high expression of YAP/YAP1 and/or TAZ remained associated with worse OS and/or DFS after adjustment, with hazard ratios indicating a clinically relevant increase in the risk of death or recurrence (HR=4.351; 95% CI: 1.477–12.819; p=0.008 for high TAZ expression32). Although Hippo pathway biomarkers were included in Cox models in other studies [44,46,47], isolated YAP1 expression did not emerge as an independent predictor of OS or DFS, despite its association with more aggressive histopathological features in univariable analyses. In addition, several studies [36,38–41] reported an association between Hippo pathway activation (or combined expression profiles, such as YAP/TAZ-activated gene signatures) and worse prognosis only in univariable or qualitative analyses, without presentation of Cox models.

In the remaining 11 studies, the assessment of the prognostic impact of Hippo pathway biomarkers was limited. Six studies [36,38–41,45] analyzed only Kaplan–Meier curves, without Cox models, generally showing worse OS, Disease-Free Survival (DFS), or Recurrence Free Survival (RFS) in patients with high expression or nuclear localization of YAP/YAP1 and/or TAZ. The other five studies [33,37,42,43,48] did not perform survival analysis, restricting their evaluation to correlations between biomarker expression and clinicopathological variables (such as stage, grade, metastases, and EMT) without time-to-event outcomes.

### Quantitative synthesis

#### Analysis of YAP

The meta-analysis of YAP expression and OS revealed a pooled HR of 1.918 (95% CI: 1.036 – 3.549) using a random-effects model. This statistically significant association (p=0.038) indicates that high YAP expression is significantly correlated with a poorer prognosis, suggesting an increased risk of mortality in patients with OSCC. The pooled effect size, was positioned entirely to the right of the null effect line (HR=1), confirming an adverse prognostic role for YAP overexpression (Figure 2).

**Fig. 2.**
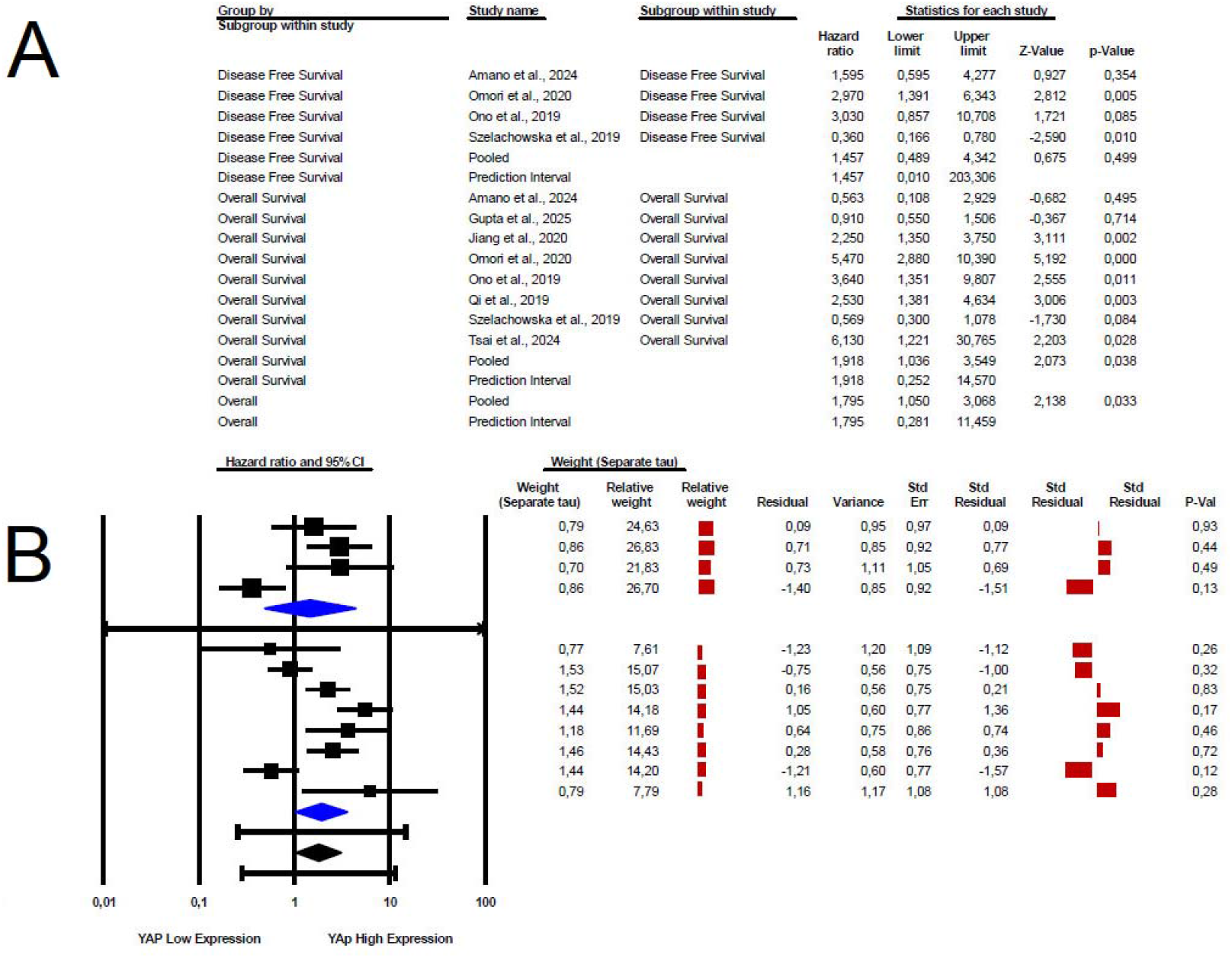
The prognostic value of YAP immunohistochemical expression in OSCC. (A) Association between high nuclear YAP expression and Overall Survival (OS). (B) Association between high nuclear YAP expression and Disease-Free Survival (DFS). Note: The squares represent the hazard ratio (HR) for each individual study, with the size proportional to the study’s weight in the meta-analysis. Horizontal lines indicate the 95% confidence interval (95% CI). The diamond at the bottom represents the pooled HR. An HR value greater than 1 indicates that high YAP expression is significantly associated with poor survival outcomes.

Substantial statistical heterogeneity was observed across the included studies. The I^2^ statistic was 82.4%, indicating a high level of inconsistency. This was further supported by a significant Chi-square test (Q=39.06, p<0.001). The between-study variance, Tau^2^, was estimated at 0.588. Despite this high heterogeneity, the direction of the effect consistently pointed towards an increased risk associated with high YAP expression across most primary studies, suggesting that the variability primarily lies in the magnitude rather than the direction of the prognostic effect. This consistency in effect direction supports the aggregation of data, albeit with appropriate caution.

The funnel plot for YAP revealed a noticeable asymmetry, characterized by a scarcity of small studies with negative or non-significant findings in the lower-left quadrant. This pattern suggests the potential presence small-study effects, where smaller studies with positive or statistically significant results are more likely to be published. However, the overall effect size (HR>2.0) remained robust, indicating a strong prognostic impact of YAP despite these potential biases (Figure 3a). The interpretation of funnel plot asymmetry should be made with caution, particularly when the number of included studies is limited.

**Fig. 3.**
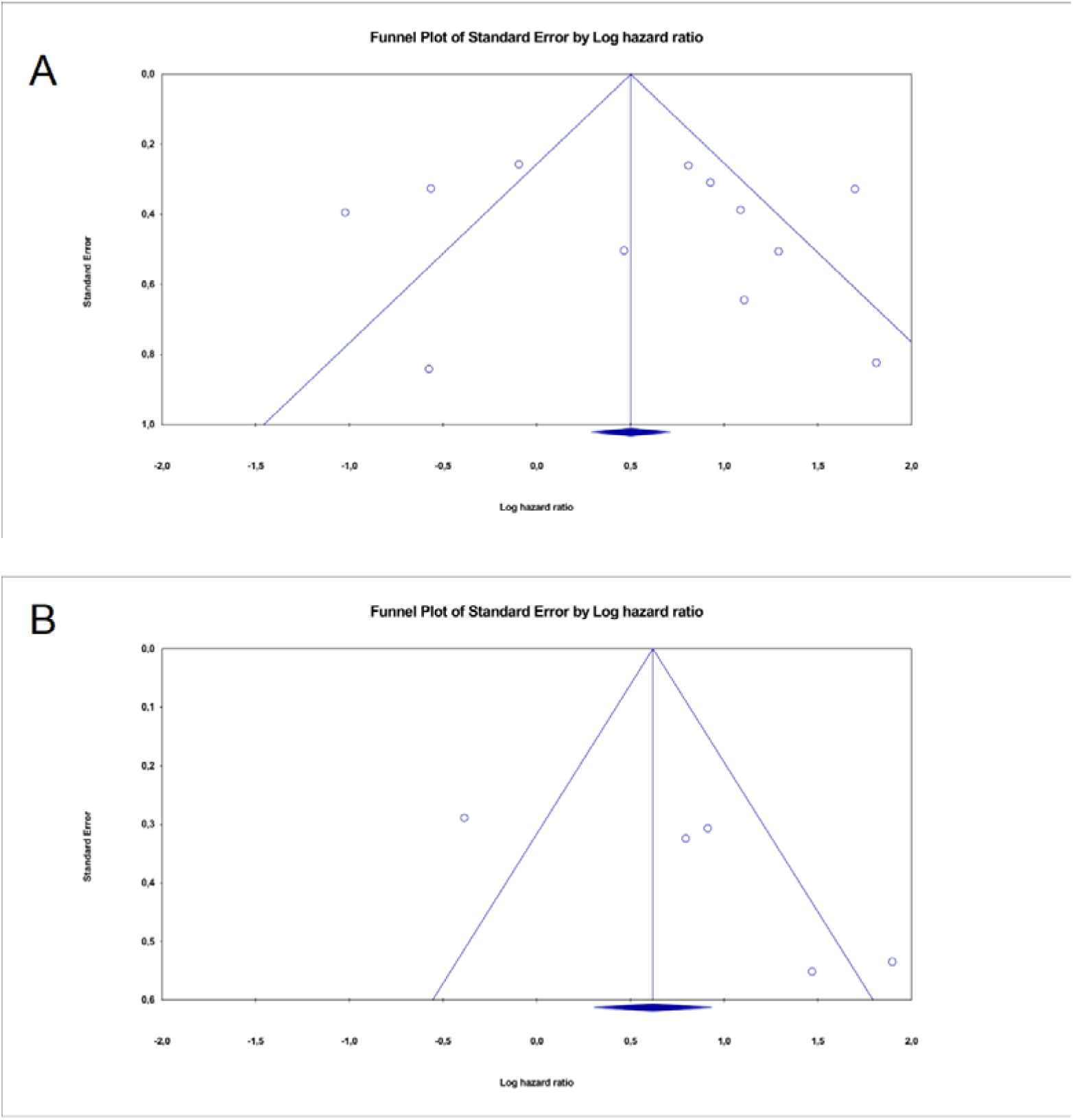
(A). Funnel plot assessing potential publication bias and heterogenity in the meta-analysis of YAP expression. This scatter plot illustrates the distribution of the included studies regarding YAP expression and survival outcomes. The horizontal axis represents the effect size (Log Hazard Ratio), while the vertical axis indicates the standard error (SE) of the estimate; (B) Funnel plot for the evaluation of publication bias in studies investigating TAZ expression. The scatter plot displays individual studies included in the meta-analysis of TAZ survival outcomes. The x-axis represents the effect size (Log Hazard Ratio), and the y-axis represents the precision of the study (Standard Error). The plot symmetry is used to assess the presence of potential publication bias or small-study effects across the analyzed studies. Fig. 4 Forest plots depicting the prognostic significance of TAZ expression in OSCC. (A) Meta-analysis of the association between high TAZ expression and Overall Survival (OS). (B) Meta-analysis of the association between high TAZ expression and Disease-Free Survival (DFS). Note: Individual study effect sizes are represented by squares, with horizontal lines indicating 95% confidence intervals (CI). The diamond represents the pooled Hazard Ratio (HR), where an HR > 1 signifies that TAZ overexpression is a risk factor for poor survival outcomes.

#### Analysis of TAZ

For the TAZ biomarker, meta-analysis yielded a pooled HR of 2.845 (95% CI: 1.682 – 4.811) for OS, calculated using a random-effects model. This result was statistically significant (p = 0.0001), indicating that elevated TAZ expression is significantly associated with a worse prognosis. The confidence interval for the pooled HR did not include unity, confirming that high TAZ expression confers an increased risk of adverse outcomes, thereby favoring the control group (low TAZ expression) in terms of survival (Figure 4).

**Fig. 4.**
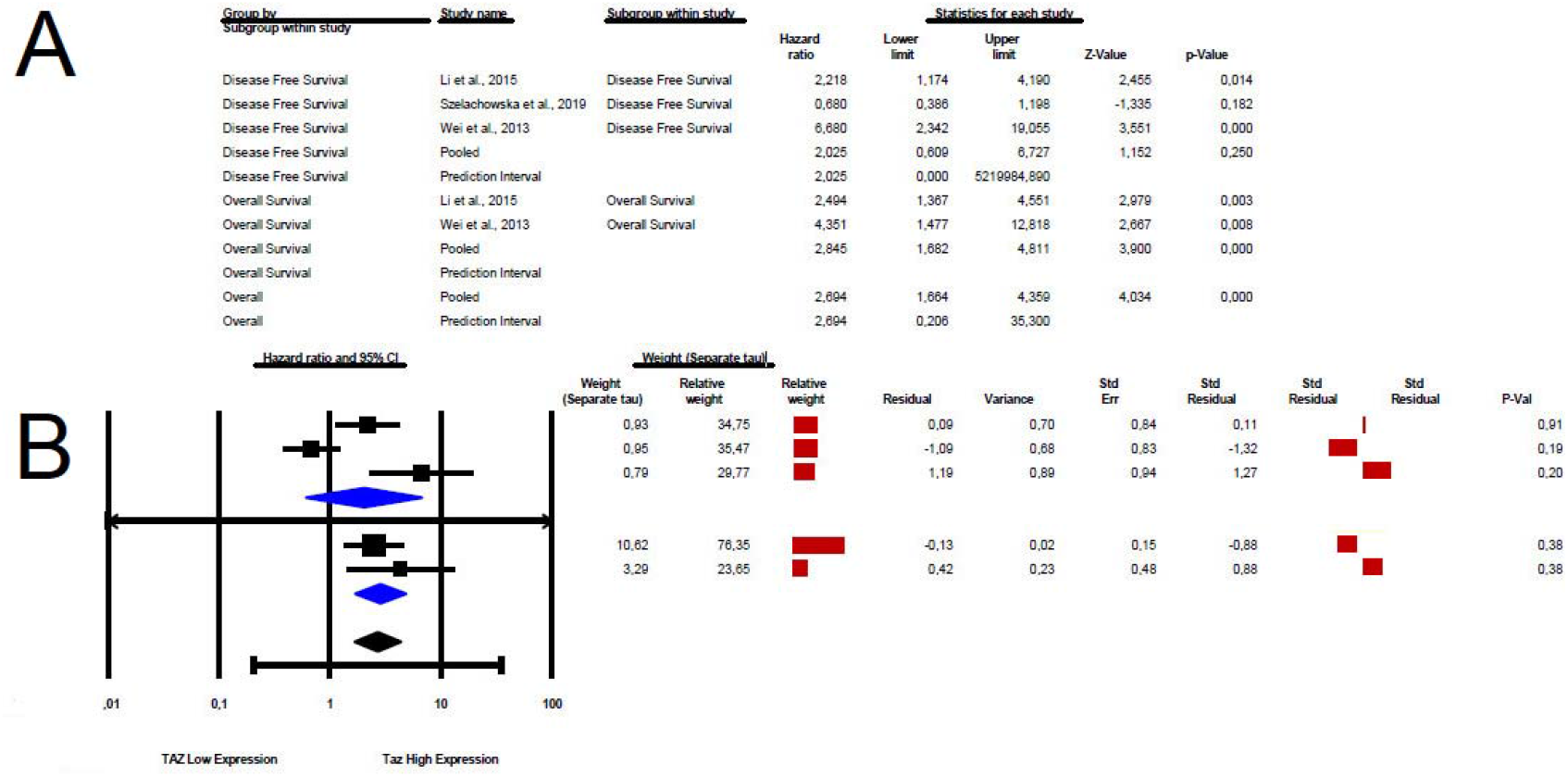
Forest plots depicting the prognostic significance of TAZ expression in OSCC. (A) Meta-analysis of the association between high TAZ expression and Overall Survival (OS). (B) Meta-analysis of the association between high TAZ expression and Disease-Free Survival (DFS). Note: Individual study effect sizes are represented by squares, with horizontal lines indicating 95% confidence intervals (CI). The diamond represents the pooled Hazard Ratio (HR), where an HR > 1 signifies that TAZ overexpression is a risk factor for poor survival outcomes.

Significant heterogeneity was detected among the studies evaluating TAZ. The I2 statistic was 81.3%, signifying a high degree of inconsistency. The Chi-square test (Q=21.436, p<0.001) further confirmed this substantial variability. The estimated between-study variance, Tau2, was 0.593, reflecting the dispersion of true effects across studies. The presence of high heterogeneity necessitated the use of a random-effects model to provide a more conservative estimate of the pooled effect and its confidence interval.

The assessment of publication bias for TAZ was challenging due to the limited number of included studies (n=3) and the high observed heterogeneity (I2=81.3%). In such scenarios, the interpretation of funnel plot asymmetry can be unreliable, as scatter may be primarily attributable to true heterogeneity rather than publication bias (Figure 3b) Furthermore, statistical tests for funnel plot asymmetry, such as Egger’s test, are typically underpowered with fewer than 10 studies, making their application less informative. While publication bias cannot be definitively ruled out, the results should be interpreted with caution, acknowledging the inherent limitations imposed by the small sample size of included studies. The observed scatter in the funnel plot is likely influenced by the high heterogeneity due to the small number of studies included as well as their size.

## DISCUSSION

OSCC remains an important global health problem, with projections of a 30–40% increase in incidence and mortality by 2040 [49]. In this context, there is an urgent need to develop and validate more accurate diagnostic and prognostic approaches, and several authors have emphasized the role of prognostic biomarkers to improve outcome prediction in OSCC and head and neck squamous cell carcinoma beyond conventional staging [50]. The identification of robust prognostic biomarkers capable of refining risk stratification beyond the traditional TNM system (tumour-node-metastasis) therefore represents a key strategy to improve patient survival [51]. In this regard, the present systematic review identified a consistent pattern of high, particularly nuclear, expression of YAP/TAZ and related markers, which was strongly associated with adverse clinicopathological features and with significantly poorer overall and DFS. In this context, the expression patterns of Hippo pathway effectors observed in OSCC appear to reflect a logic in which mechanical and oncogenic signals converge on YAP/TAZ activation. In our systematic review, most studies reported high expression of YAP/YAP1 and/or TAZ in OSCC, predominantly with nuclear or nucleocytoplasmic staining. In addition, the data also suggested attenuation of upstream inhibitory components, such as low MST1/LATS1/LATS2 expression alongside YAP1 positivity, or regulatory events consistent with increased YAP/TAZ activity, such as LATS2 downregulation (Table 4). This OSCC pattern is aligned with evidence that YAP/TAZ hyperactivation is frequent in solid tumors and contributes to proliferation, EMT, invasion, and metastasis [52]. Likewise, studies on hepatocellular carcinoma [53] and renal cancers [54] have associated YAP/TAZ overexpression/nuclear expression with advanced disease and poor prognosis, and recent clinical work has even proposed YAP-based prognostic models to refine risk prediction beyond standard parameters. However, the breast cancer literature [55] indicates that YAP may be associated with poor prognosis in certain subtypes, whereas in ER-positive tumors this relationship may differ or even be reversed, suggesting that the clinical impact of YAP/TAZ is not uniform and may depend on the biological context of the subtype, lineage-specific transcriptional architecture, and treatment-imposed selective pressures. In this context, our OSCC findings, predominantly YAP/TAZ overexpression with concomitant signals of weakened upstream inhibition, fit the prevailing oncogenic model of Hippo pathway deregulation.

**Table 4.**
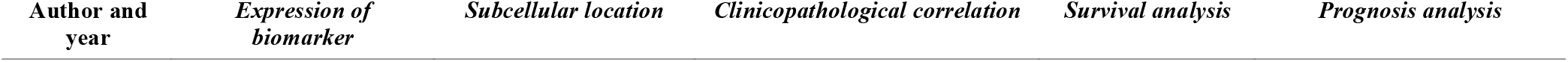
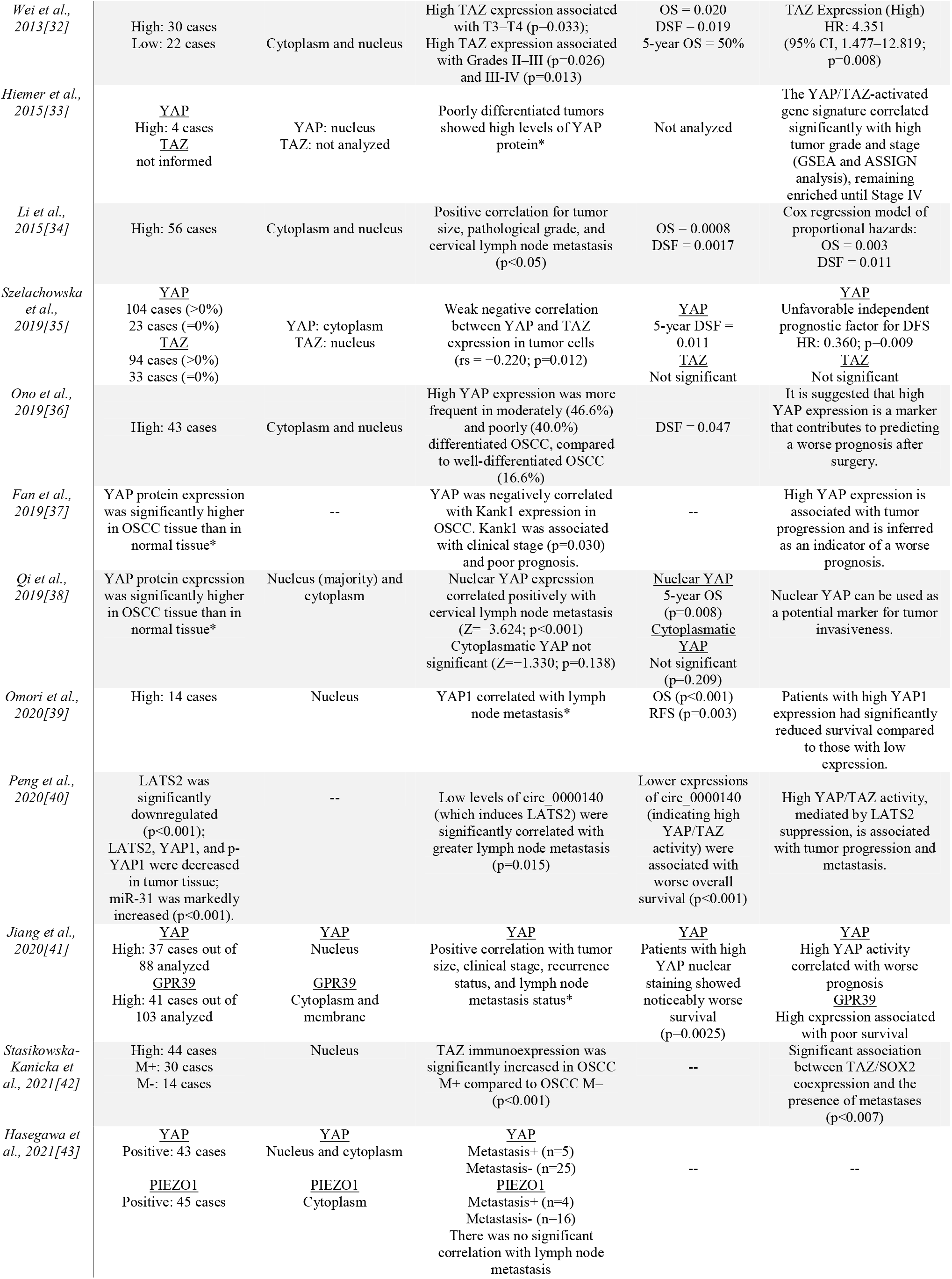
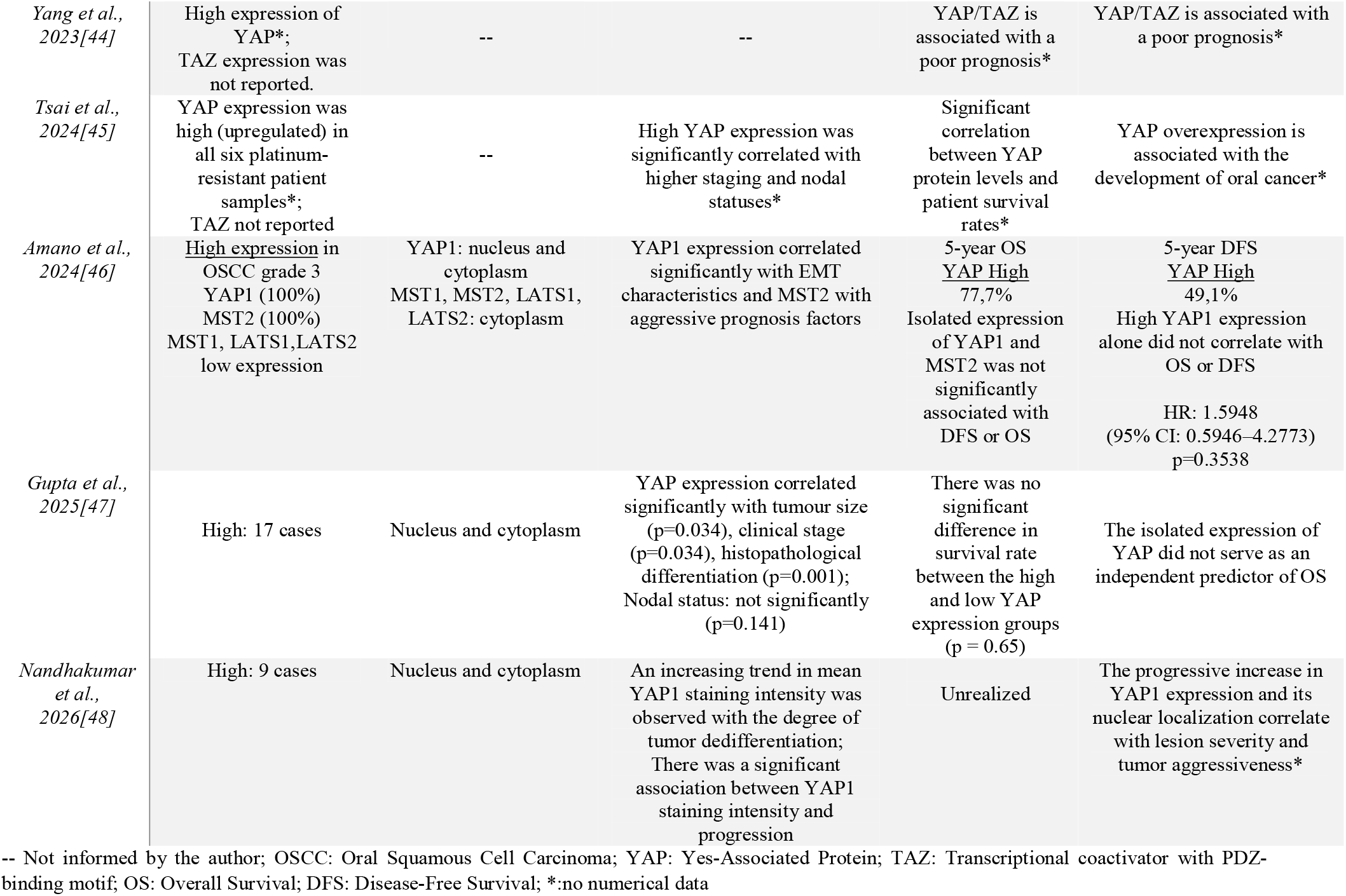
Summary of the main results of the eligible studies.

In addition to the level of YAP/TAZ expression, their subcellular localization also has biological and clinical relevance. In general, activation of the Hippo pathway culminates in the phosphorylation of YAP/TAZ by MST1/2 and, especially, LATS1/2, which favors interactions with regulatory proteins and promotes their cytoplasmic retention/degradation, being associated with a functionally inactive state. However, nuclear accumulation of YAP/TAZ indicates relative dephosphorylation and TEAD-mediated transcriptional activation, with the potential induction of pro-oncogenic programs [33,56]. Similarly, in renal [57,58] and hepatocellular tumors [59], alterations in polarity and cell-adhesion proteins have been implicated in facilitating YAP/TAZ nuclear translocation and enhancing invasive and metastatic phenotypes. In renal carcinoma, studies [58] indicate that YAP/TAZ activation (including nuclear enrichment of TAZ) is associated with higher stage/grade and poorer survival, and that polarity regulators such as Par3 may act as pro-metastatic cofactors via the YAP/TAZ axis. In hepatocellular carcinoma65, loss of apical polarity determinants such as MPP5/PALS1 correlates with nuclear accumulation of YAP/TAZ and worse outcomes. In the present review, nuclear or nucleocytoplasmic staining of YAP/YAP1 and TAZ was the dominant pattern in OSCC, reported in most studies, and this pattern was linked to parameters of aggressiveness and poorer outcomes, consistent with observations in other anatomical sites.

Despite the consistent direction of effect, our meta-analysis revealed substantial statistical heterogeneity among the included studies for both YAP (I^2^ = 82.4%) and TAZ (I^2^ = 81.3%). This high level of heterogeneity, while common in meta-analyses of biomarker studies, necessitates careful interpretation. Several factors likely contribute to this variability. Differences in immunohistochemical (IHC) staining protocols, including antibody clones, dilution, and detection systems, can significantly impact the assessment of protein expression. Moreover, the lack of standardized cut-off values for defining “high” versus “low” expression across studies introduces considerable variability. Some studies used median values, others used quartiles, and some employed receiver operating characteristic (ROC) curve analysis, leading to diverse thresholds. Furthermore, variations in patient populations (e.g., geographical origin, ethnicity), tumor characteristics (e.g., stage, differentiation grade, anatomical subsite), and follow-up durations among the primary studies could also contribute to the observed heterogeneity. Analysis of the funnel plot for YAP suggested some asymmetry, with a potential underrepresentation of small studies reporting non-significant or negative findings. However this “small-study effect” could lead to an overestimation of the true effect size. For TAZ, the limited number of studies (n=3) restricts the power of visual or statistical tests for publication bias, and the observed scatter in the funnel plot is more likely attributable to the high heterogeneity rather than clear evidence of bias. While these potential biases are acknowledged limitations, the magnitude and statistical significance of the pooled HRs for both YAP and TAZ, particularly for YAP, suggest that their prognostic utility is unlikely to be entirely spurious.

This meta-analysis, while comprehensive, is subject to several limitations. Firstly, the high degree of heterogeneity observed for both YAP and TAZ, primarily due to methodological differences in biomarker assessment and diverse patient cohorts, warrants cautious interpretation. Although a random-effects model was employed to account for this variability, residual confounding cannot be entirely excluded. Secondly, the relatively small number of studies included for TAZ (n=3) limits the robustness of its pooled estimate and the power to detect publication bias. Thirdly, the reliance on immunohistochemistry for biomarker assessment, while common, introduces inherent variability and subjectivity. Future studies should consider more quantitative methods, such as Reverse Transcription Polymerase Chain Reaction (qRT-PCR) or Western blot, or explore the prognostic value of YAP/TAZ activation status rather than just total protein expression.

Survival analysis represents a fundamental step in translating this biological evidence into prognostic utility, allowing evaluation of whether YAP/TAZ activation effectively translates into an increased risk of death and/or recurrence [51]. In the present systematic review, 12 of the 17 studies performed time-to-event analyses, and six applied Cox regression. In most studies, high expression of YAP/YAP1 and/or TAZ, particularly when nuclear, was associated with poorer OS and/or shorter DFS/RFS, and in at least three studies [32,34,35] the effect remained independent after adjustment for clinicopathological variables (for example, high TAZ: HR=4.351; 95% CI: 1.477–12.819; p=0.008).

Across multiple solid tumors, activation of the YAP/TAZ axis has been associated with tumor proliferation, invasion, metastasis, and poorer survival, with evidence supporting its prognostic value in human cohorts [60]. In pancreatic ductal adenocarcinoma, studies [61] describe YAP/TAZ as mediators of progression and resistance, frequently associated with unfavorable prognosis in clinical series and syntheses of the field. Similarly, clinical evidence [62] indicates that YAP activation is linked to more aggressive gastric cancer subtypes and poorer outcomes, reinforcing the view of YAP/TAZ as amplifiers of tumor progression in digestive tract epithelia. In non-small cell lung cancer, focused reviews [63] on YAP/TAZ also describe associations with adverse clinical evolution and suggest potential prognostic/predictive utility in specific therapeutic contexts. Herein, we reinforce that Hippo pathway deregulation and YAP/TAZ activation, particularly with a nuclear pattern, constitute a biologically and clinically relevant axis in OSCC progression, being consistently associated with tumor aggressiveness and adverse outcomes. Nevertheless, it is important to emphasize that not all cohorts across different tumors demonstrate prognostic independence for YAP/TAZ, suggesting a context-dependent role influenced by molecular subtype, microenvironment, and co-activated pathways.

The findings from our meta-analysis position YAP and TAZ as promising prognostic biomarkers for stratification of OSCC patients. In a clinical setting, identifying patients with high YAP/TAZ expression could help tailor more aggressive treatment strategies, including intensified adjuvant therapy or closer surveillance. Conversely, patients with low expression might be spared unnecessary aggressive treatments. However, for these biomarkers to be adopted into routine clinical practice, standardization of IHC protocols and definition of robust, clinically validated cut-off values are imperative. The development of a standardized scoring system would enhance reproducibility and comparability across institutions for the development of additional multicenter prospective studies.

## CONCLUSION

Overexpression of Hippo pathway biomarkers, particularly YAP/YAP1 and/or TAZ, is associated with greater tumor aggressiveness and progression and poorer survival in patients with OSCC.

## Data Availability

All data produced in the present study are available upon reasonable request to the authors

## DECLARATIONS

### Ethics approval and consent to participate

not applicable

### Consent for publication

not applicable.

### Human Ethics and Consent to Participate declarations

not applicable

### Consent for publication

not applicable

## Acknowledgements

The authors acknowledge funding from Conselho Nacional de Desenvolvimento Científico e Tecnológico (406258-2022-8 to M.L.L) and the U.S. National Institutes of Health (R01CA206880 to A.J.E. and R01CA280279 to A.J.E. and M.L.L.) and a cy pres research award from the Krueger v. Wyeth settlement fund (to A.J.E.).

## Authors’ contributions

Igor Felipe Pereira Lima: Conceptualization, Methodology, Investigation, Data curation, Formal analysis, Software, Writing – original draft, Writing – review & editing.

Petra Gonçalves Reinehr: Investigation, Methodology, Writing – original draft.

Leonardo da Silva Bittencourt: Data curation, Formal analysis, Methodology, Software, Writing – review & editing.

Adam Jeffrey Engler: Funding acquisition, Investigation, Visualization, Writing – review & editing.

Marcelo Lazzaron Lamers: Conceptualization, Investigation, Supervision, Project administration, Funding acquisition, Writing – review & editing.

